# A Post-Marketing Evaluation of Avacopan Safety with a Focus on Hepatic Adverse Events

**DOI:** 10.64898/2026.07.20.26358315

**Authors:** Punam Bharania, Michelle Geller, Atasi Jana, Kelsey Mirkovic, Zachary S. Wallace, Alana Bozeman, Sunil Mehta, Byeong Yoon, Paul Burton

**Affiliations:** Amgen Limited, Uxbridge, United Kingdom; Amgen, Inc., 1 Amgen Center Drive, Thousand Oaks, California, USA, 91320

**Keywords:** avacopan, ANCA-associated vasculitis, drug-induced liver injury, hepatotoxicity, vanishing bile duct syndrome, pharmacovigilance

## Abstract

**Background:** Avacopan, an oral complement C5a receptor inhibitor, has been shown to help patients with anti-neutrophil cytoplasmic antibody (ANCA)-associated vasculitis (AAV) achieve and sustain remission with reduced glucocorticoid exposure and associated toxicity. As of January 20, 2026, estimated global post-marketing exposure exceeded 25,000 patient-years. Hepatic adverse events (AEs), predominantly liver enzyme elevations observed during clinical development, remain an important post approval safety risk. Since approval, vanishing bile duct syndrome (VBDS), a rare but serious complication of drug induced liver injury (DILI) has been reported with avacopan use. This analysis evaluated Amgen’s global safety database data to characterize hepatic adverse event reports associated with avacopan, including VBDS.

**Methods:** We conducted a retrospective descriptive analysis of hepatic AEs recorded in the Amgen global safety database.

**Results:** As of 20 January 2026, serious hepatic AEs were reported at approximately 31 events/1,000 patient-years. The majority of hepatic events comprised laboratory abnormalities that resolved following avacopan discontinuation. Thirty-one VBDS cases were reported and, of those, 27 originated from Japan. Fatal VBDS cases were reported predominantly from Japan and occurred in patients >65 years.

**Conclusions:** Hepatotoxicity remains an important avacopan-specific safety risk. Post-marketing data indicate that most hepatic events are reversible laboratory abnormalities; however, serious liver injury and VBDS, including fatal outcomes, have been reported, predominantly from Japan. No new safety risks have emerged from the ongoing Japanese post-marketing study. Further investigation is warranted to understand potential mechanisms underlying the disproportionate occurrence of serious VBDS events in Japanese patients and inform optimized risk-mitigation strategies.

## INTRODUCTION

Avacopan is an orally administered selective complement C5a receptor inhibitor used with standard immunosuppressive therapy for anti-neutrophil cytoplasmic antibody (ANCA)-associated vasculitis (AAV), including granulomatosis with polyangiitis (GPA) and microscopic polyangiitis (MPA).^1,2^ AAV is a rare, serious, immune-mediated vasculitis that can cause major organ damage, including end-stage kidney disease, and can be life-threatening. Prior to the introduction of immunosuppressive therapy, outcomes for patients with GPA and MPA were poor, with more than 80% of patients dying within the first year following diagnosis.^3^ Despite substantial advances in treatment, mortality among patients with GPA and MPA remains significantly elevated, with studies reporting a 2.6-fold or greater increased risk of death compared with the general population.^4,5^ In addition to increased mortality, GPA and MPA are associated with relapse, treatment-related toxicities, particularly from glucocorticoids, and substantial impairments in health-related quality of life (HRQoL).^6–10^

Evidence from clinical trials and real-world studies demonstrate that patients treated with avacopan achieve and sustain remission at high rates while reducing glucocorticoid exposure and associated toxicity.^11–13^ Collectively, these studies also support the impact of avacopan on lowering the risk of relapse as well as improving kidney recovery and quality of life.^12,14–18^ Based on evidence from the clinical trial program, avacopan received its first approval in Japan in September 2021, followed by approvals in the United States, European Union, and other regions.^1,19,20^ International guidelines for the management of AAV also recommend the use of avacopan, especially as part of a regimen to minimize glucocorticoid exposure and improve kidney outcomes.^21,22^ Although randomized, controlled clinical trials and real-world observational studies have demonstrated the safety of avacopan, the accumulating post-approval experience provides an opportunity to further characterize the safety profile of avacopan beyond these studies.

Hepatotoxicity was observed during avacopan clinical development and, since initial approvals, has been described as a known safety risk in product labeling with guidance on risk mitigation including hepatic monitoring and discontinuation.^19,20,23^ In the pivotal ADVOCATE trial, serious hepatic events occurred at a rate of 5.4%. These events were mainly hepatic laboratory abnormalities and resolved after withdrawal of avacopan and/or other potentially hepatotoxic medicinal products.^20^ Serious hepatic events have also occurred in the post-marketing setting. A type of serious hepatic event, vanishing bile duct syndrome (VBDS), has also been reported.

VBDS is a rare but serious complication of drug induced liver injury (DILI) characterized by the progressive destruction and disappearance of the small intrahepatic bile ducts.^24,25^ This destruction leads to loss of bile ducts and chronic cholestasis, which can ultimately cause severe liver damage and failure. VBDS is diagnosed on liver biopsy which shows a loss of intrahepatic bile ducts in greater than 50% of portal areas, provided 10 or more portal tracts are evaluated.

Some hepatic events, including VBDS, have had fatal outcomes.^26,27^

Structured hepatic monitoring is an important risk mitigation activity for avacopan use. This need is not unique to avacopan. The FDA Drug-Induced Liver Injury Rank (DILIrank) 2.0 dataset includes 1,336 FDA-approved drugs and categorizes 217 as very-most-DILI-concern drugs, illustrating the broader clinical importance of proactive liver-risk mitigation for approved therapies.^28^ Nevertheless, serious hepatic events and the emergence of VBDS in association with avacopan use warrants careful evaluation.

This report summarizes post-approval evidence regarding the hepatic safety of avacopan, with emphasis on serious hepatic abnormalities and events including VBDS and discusses risk-mitigation measures that are important to minimize the risk of serious hepatic events.

## METHODS

The Amgen global safety database, which includes all post-approval adverse events reported to Amgen as well as serious adverse events from clinical trials, was used as the source database for this analysis. Reports included those received globally from healthcare professionals, post-marketing studies, patients, and other spontaneous sources, as well as cases identified from the published literature. The spontaneous reports received from Healthcare Providers (HCPs) can have limited information. Although Amgen applies its usual process for following up with the reporter to provide further information; data may ultimately remain missing. All events meeting the predefined search criteria were retrieved up to 20 January 2026. All information available in the adverse event report was reviewed. The predefined search criteria were drug-related hepatic disorders, comprehensive search standard MedDRA query (SMQ) and biliary tract disorders SMQ (broad) MedDRA v28.1 for all cases (both serious and non-serious). Serious adverse events were defined per the ICH guidelines.^29^

The patient exposure calculation for avacopan assumes that the average administration is the recommended dosage of 30 mg BID for all indications. Avacopan is supplied in 10 mg capsules for oral administration. Therefore, 6 capsules (60 mg) correspond to 1 patient day. Estimated exposure (in patient-years) is calculated as follows:

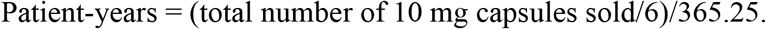

## RESULTS

As of January 20, 2026, cumulative post-marketing exposure to avacopan is estimated to exceed 25,000 patient-years, including 6,511 patient-years in the United States and 19,240 patient-years in other marketed countries. Japan accounted for 11,679 patient-years, approximately 45% of worldwide estimated exposure.

Data from the Amgen global safety database shows that most cases of hepatic events report aminotransferase or liver-function abnormalities that appear monitorable and reversible after avacopan interruption or discontinuation. The five most reported events to January 20, 2026, were abnormal hepatic function (n=337), liver disorder (n=199), increased hepatic enzymes (n=156), drug-induced liver injury (n=82), and increased liver function tests (n=69). Among cases with available time to onset information, time to onset in days from the first dose of avacopan had a mean of 72.55 days and median of 51 days. Serious hepatic adverse events were reported at a rate of approximately 31 events per 1,000 patient-years. The majority of serious hepatic events, including fatal cases, were predominantly reported from Japan, often in elderly female patients (> 65 years old). A causal relationship to avacopan could not be confirmed in all cases because of incomplete clinical information, concomitant medications (e.g. sulfamethoxazole-trimethoprim), and competing etiologies (e.g., concurrent infections, septic shock, and multiorgan failure).

As of January 20, 2026, 31 cases of VBDS (which are included in the total number of hepatic events described above) were reported, consisting of 27 from Japan, 2 from Canada (1 patient of Chinese race and 1 patient of unknown race), and 2 cases in patients of unknown race from literature reports from China that used the US Food and Drug Administration Adverse Event Reporting System (FAERS) database. No cases of VBDS were reported from the European Union or United States. Where reported (n = 16), time to VBDS onset ranged from 22 to 90 days. Of the 31 reported VBDS cases, 15 were confirmed by liver biopsy.

Among the biopsy-confirmed cases, 9 were fatal, 1 had not recovered, 1 had recovered, and 4 were recovering at last follow-up. Sulfamethoxazole/trimethoprim was a co-suspect drug in 8 biopsy-confirmed cases, and six fatal cases had relevant clinical confounders (e.g., concurrent infection, septic shock and multi organ failure) that prevent conclusive attribution to avacopan. All fatal VBDS cases were reported from Japan in patients older than 65 years.

## DISCUSSION

In clinical trials, the benefits of avacopan, including high rates of remission and sustained remission with significantly reduced glucocorticoid exposure, were observed in the context of hepatic adverse events. Thus, hepatotoxicity was identified as a risk with the use of avacopan and included in labels globally.^30^

The DILIrank 2.0 dataset includes several medications used across multiple specialties for immune mediated, inflammatory or related conditions including azathioprine, methotrexate, leflunomide, sulfasalazine, infliximab, allopurinol, and febuxostat. The dataset further includes antibiotics frequently prescribed across healthcare specialties, such as fluoroquinolones, as well as medications used for common chronic conditions, including hypertension and other cardiovascular disorders, such as carvedilol and diltiazem, and mood disorders, such as duloxetine. Acetaminophen, one of the most widely used over-the-counter medications, is also included, underscoring that liver-risk monitoring and mitigation are broadly relevant across commonly used therapeutic classes.

Most cases of hepatic events among avacopan users involved aminotransferase elevations or liver-function abnormalities that were reversible after avacopan interruption or discontinuation; however, VBDS has emerged as a newly identified manifestation of severe liver injury among avacopan users. The occurrence of VBDS, including events with fatal outcomes, indicates that early recognition and decisive management are essential.

Reports from Japan account for a disproportionate share of serious hepatic events and nearly all VBDS cases. Potential explanations may include differences in patient characteristics, regional labeling variation based on health authority label language requirements (**Table 1**), treatment practices, concomitant medications, ascertainment and reporting, background disease phenotype, and pharmacogenetic susceptibility.

**Table 1.** Hepatic-safety labeling and monitoring comparison for avacopan in the United States and Japan as of 20 January 2026*.

| Domain | U.S. Prescribing Information (USPI) for TAVNEOS | Japan Prescribing Information (JPI) for TAVNEOS | EU Summary of product characteristics (SmPC) |
| --- | --- | --- | --- |
| **Warning and precautions** | Named hepatotoxicity warning. Obtain ALT, AST, ALP, and total bilirubin before treatment; repeat every 4 weeks for 6 months, then as clinically indicated. | Important Basic Precautions: hepatic dysfunction may occur; perform liver-function tests before and periodically during treatment. | Named hepatotoxicity warning, including VBDS seen in the post marketing setting. Obtain ALT, AST, ALP, and total bilirubin before treatment; repeat every 4 weeks for 6 months, then as clinically indicated. |
| **Management of abnormal liver tests** | ALT/AST >3x ULN: evaluate promptly and consider interruption. ALT/AST >5x ULN, or >3x ULN with bilirubin >2x ULN: discontinue until DILI is ruled out. | If an abnormality is observed, discontinue treatment or take other appropriate measures. | ALT/AST >3x ULN: Treatment must be re-assessed clinically and temporarily stopped<br>ALT/AST >5x ULN, or >3x ULN with bilirubin >2x ULN: discontinue until DILI is ruled out. |

\*The EU SmPC, JPI and USPI have subsequently been updated with additional monitoring.
Abbreviations: ALP, alkaline phosphatase; ALT, alanine aminotransferase; AST, aspartate aminotransferase; DILI, drug-induced liver injury; PI, prescribing information/package insert; ULN, upper limit of normal; VBDS, vanishing bile duct syndrome.

Avacopan is primarily metabolized by CYP3A4.^31^ Genetic background may play a significant role in susceptibility to avacopan-induced liver injury. The CYP3A5*3 allele is highly prevalent in the Japanese population (allele frequency, approximately 85%), and the *3/*3 homozygous genotype is estimated to be present at a frequency of approximately 72%.^32^ CYP3A4 and CYP3A5 are highly homologous drug-metabolizing enzymes; however, their functional roles differ. In Japanese individuals, the CYP3A5*3 allele is common and leads to the loss of CYP3A5 activity, making CYP3A4 the primary enzyme responsible for hepatic drug metabolism.^32,33^ Some CYP3A polymorphisms unique to the Japanese population can further reduce the CYP3A4 function, which may increase susceptibility to avacopan-related adverse effects. This pharmacogenetic hypothesis is plausible but remains unproven.

Additional genetic factors may also contribute to susceptibility. In Japanese patients, MPO-ANCA-positive microscopic polyangiitis is strongly associated with HLA-DRB1*09:01, and Tsuchiya and colleagues identified the HLA-DRB1*09:01-DQB1*03:03 haplotype as a principal HLA-region genetic risk factor for microscopic polyangiitis.^34^ HLA-DRB1*09:01 has also been reported as a genetic marker associated with susceptibility to, and more severe progression of, primary biliary cholangitis, a cholestatic liver disease that may share clinical features with or mimics VBDS. Whether antigen presentation related to this haplotype contributes to susceptibility to avacopan-associated VBDS remains speculative and requires further investigation.

Notably, at the time of the data cutoff, regional labeling differed (**Table 1**), and risk-management language has evolved as post-marketing evidence has accumulated. Since product approval, the United States prescribing information has included a warning for hepatotoxicity that could be life-threatening and recommends baseline testing of alanine aminotransferase, aspartate aminotransferase, alkaline phosphatase, and total bilirubin. It also recommends repeat monitoring every 4 weeks for the first 6 months and as clinically indicated thereafter; it specifies criteria for avacopan treatment interruption and discontinuation.^19^ The European product information: (i.) describes post-marketing DILI and VBDS, including fatal outcomes; (ii) recommends baseline and repeated liver monitoring; and (iii.) has criteria for treatment interruption and discontinuation.^20^ Recent FDA and PMDA/MHLW safety communications emphasize the importance of intensive, early monitoring of liver function tests and prompt discontinuation when clinically significant abnormalities or cholestatic symptoms occur.^26,27^ Of note, the European, Japan, US and additional labels have been recently updated to include additional monitoring in patients and further discontinuation recommendations.^2,19,35^

In addition to spontaneous reports received, the interim safety analysis based on the ongoing Japanese post-marketing surveillance included 451 patients with a data cutoff of September 26, 2025, of whom 97 patients (21.5%) experienced hepatic function disorder adverse drug reactions during the first 12 months, of which 24 were considered serious (5.3%), including 3 DILI cases (0.7%), and 1 VBDS case (0.2%). Most events occurred early: 90% within 112 days and 54% within 56 days.^36^

Since the approval of avacopan, this accumulating post-approval experience provides an opportunity to characterize the safety profile of avacopan beyond controlled clinical trials. However, an inherent limitation of this review is the nature of the information provided in these spontaneous reports, including underreporting, variable completeness, potential duplicate reporting, incomplete follow-up, and confounding by concomitant therapies and underlying disease. Therefore, event counts should not be interpreted as incidence rates unless exposure-adjusted denominators are specified.

## CONCLUSION

AAV is a rare, serious, immune-mediated vasculitis that can cause major organ damage and life-threatening disease. Despite improvements in treatment, mortality and long-term morbidity remain substantial. Avacopan remains an important treatment option for appropriate patients because of its demonstrated benefits, including high rates of remission and sustained remission, improvements in kidney function and quality of life, and reduced glucocorticoid exposure.

Hepatotoxicity is a known and important avacopan-specific safety risk monitored through post-marketing surveillance. Serious liver injury and rare VBDS, including fatal outcomes, have been reported, with a concentration of cases in Japan and early onset after treatment initiation. These findings support liver-panel laboratory monitoring before and during treatment, heightened vigilance during the first 3 months of therapy, when most events have occurred, and prompt discontinuation of avacopan when clinically significant hepatotoxicity or VBDS is suspected. Ongoing pharmacovigilance and post-authorization safety studies are needed to better characterize risk factors, investigate potential mechanisms underlying the disproportionate occurrence of serious VBDS events in Japanese patients, and further optimize risk-mitigation strategies.

## DATA AVAILABILITY

Data underlying post-marketing pharmacovigilance summaries are derived from safety databases and regulatory sources and may not be publicly available because of privacy, pharmacovigilance, and regulatory constraints. Public regulatory documents and literature sources are listed in the References. Qualified researchers may request data from Amgen clinical studies. Complete details are available at http://www.amgen.com/datasharing

## AUTHOR CONTRIBUTIONS

Punam Bharania, Zachary S. Wallace, and Paul Burton contributed to conceptualization. Atasi Jana, Kelsey Mirkovic, Zachary S. Wallace, and Alana Bozeman provided resources. Michelle Geller, Byeong Yoon, and Paul Burton provided supervision. Punam Bharania wrote the original draft. All authors were involved in reviewing the draft or revising it critically for important intellectual content. All authors have read and approved the final version to be published.

